# Neighborhood level factors and use of cigarettes, cannabis and e-cigarettes: a population-based study among Canadian adults

**DOI:** 10.1101/2025.02.14.25322276

**Authors:** Truman Fraser Wood, Trevor JB Dummer, Cheryl E Peters, Rachel A Murphy

**Affiliations:** School of Population and Public Health, University of British Columbia, Vancouver, BC, Canada; Prevention and Health Promotion, BC Centre for Disease Control, Vancouver, BC, Canada; Prevention, Screening and Hereditary Cancer, BC Cancer, Vancouver, BC, Canada; Cancer Control Research, BC Cancer, Vancouver, BC, Canada

## Abstract

Despite public health efforts, use of legal substances such as cigarettes, cannabis and e-cigarettes are common in Canada. Most policies focus on individual level factors, which do not account for possible influences of the environments to which people belong (i.e. neighborhoods). This study aimed to identify neighborhood-level risk factors for use of cigarettes, cannabis and e-cigarettes in the Canadian Partnership for Tomorrow’s Health cohort. Participants completed questionnaires on demographics and health behaviors including recent (30-d) use of cigarettes, cannabis and e-cigarettes. Geospatial neighborhood-level measures: deprivation, gentrification, household security, labor force participation, immigration and visible minority proportion were linked via postal codes. Regression models were built to understand associations between substance use and neighborhood factors. Neighborhood material deprivation, social deprivation, and household insecurity were positively associated with odds of using cigarettes, cannabis, and e-cigarettes. Odds of using these substances was higher for participants living in gentrified neighborhoods. Lower odds of cigarette, cannabis, and e-cigarette use were found for participants living in neighborhoods with a high proportion of recent immigrants and/or visible minorities. Evidence from this study suggests interventions aimed at reducing or preventing substance use should be multidimensional, encompassing strategies directed at both individuals and neighborhoods.

## Introduction

Substance use; cigarette, cannabis, and electronic cigarette (e-cigarette) use is common in Canada [1,2], despite sustained public health efforts. Cigarette smoking has been a significant public health concern in Canada for decades, although, cigarette smoking has been trending downwards in Canada since the 1980s [3]. In 2021, there were 1.5 million less cigarette smokers in Canada compared to 2015 [4] and 10.2% of Canadian adults (age 15 and older) reported currently smoking cigarettes [2]. Cigarette smoking remains higher among individuals with low household incomes, less education, and those who are single [5]. Legal cannabis sales began in 2019 following federal legislation that permitted its sale at licensed retailers in Canada [6]. In 2020, 27% of Canadian adults (age 16 and older) reported using cannabis in the past 12 months [7]. Prevalence of past-year cannabis use was highest among young adults, about 50%, compared to 16–19-year-olds, 44%, and adults 25 years and older, about 23% [7]. E-cigarettes, also referred to as vapes and electronic nicotine delivery systems, entered the Canadian market in 2004 and are therefore a relatively novel form of substance use. In 2021, 17% of Canadian adults re-ported ever use of e-cigarette products [2]. E-cigarette use is becoming increasingly prevalent in Canada [2]. Among young adults aged 20-24, the rate of past 30-day use has increased from 13% to 17% from 2019 to 2021 [2]. This could be due to more people transitioning from using cigarettes to e-cigarettes.

Among many other health conditions, cigarette smoking is associated with the development of cancers, heart disease, and death from common illnesses (such as pneumonia) [8,9]. An estimated 72% of lung cancer cases are attributable to cigarette smoking [10]. In 2017, it was estimated that 48,000 deaths were attributable to tobacco use in Canada [8]. Our understanding of the health impacts of using cannabis and e-cigarettes is limited by the more recent legalization and introduction of these substances in Canada. Studies on cannabis and health have been limited by its illicit status in most countries, which hinders research efforts as there is hesitancy for people to report use [11]. Smoking cannabis may be linked to carbon monoxide intoxication and some studies have found associations with adverse cardiac outcomes [11]. Changing laws and social norms around cannabis use may allow clarification around these associations in the future. Under-standing factors that are associated with the use of these substances in Canada has important implications for the primary prevention of numerous chronic diseases. However, research on risk factors for substance use has historically focused on individual-level factors, such as sex or income. More recently, there has been in shift in our understanding of what can influence health and behaviors to include components of the environment where a person lives, works, and/or engages in recreation.

The social ecological model (SEM) is a framework for prevention that conceptualizes four-levels; individual, interpersonal relationships, communities, and societal (i.e. policies) [12]. The framework posits that factors belonging to one level can influence factors at all other levels of the framework [12]. The SEM can be applied to better understand substance use. Using this systems approach, individuals can influence and be influenced by their interactions with others, their community, and available resources or institutions [12]. The community level of the SEM framework includes the settings where health behaviors (i.e. substance use) may occur and characteristics of one’s environment that may influence health behaviors. Application of this framework may help to expand our understanding of risk factors for substance use and how multiple factors at the individual and built environment level may disadvantage certain people.

In reference to the SEM framework, there are several theories detailing mechanisms through which an individual’s behavior can be influenced by their neighborhood environment. The social interactive theory proposes that an individual’s social network is affected by the neighborhood they live in, ultimately impacting behaviors by influencing one’s attitudes and beliefs (including those about social norms) as well as their access to supports, information, and resources [12]. Having higher social or community cohesion is also thought to promote healthier behaviors by improving access to health information and services [13]. The structural hypothesis suggests that living in a particular neighborhood determines residents’ accessibility to certain structural environments (sidewalks, parks, etc.), institutions and businesses (community centers, grocery stores, liquor re-tailers, etc.), and employment opportunities [14]. Living in a neighborhood with variable access to the aforementioned structures may affect residents use of resources and any directly or indirectly related behaviors [14]. There is also evidence that the neighborhood one lives in can impact their exposure to stressors, potentially related to their neighborhood social environment and their access to specific resources [15]. Stress can have strong impacts on behavior, especially when stress originates from an unavoidable source such as the area where one lives [15].

There is very limited research on how living in certain neighborhood is associated with substance use. However, there is some evidence to support that the forementioned theories can be applied to substance use behaviors. For example, there is evidence that social motives and social norms influence substance use behaviors [16,17]. Additionally, there is evidence that substance use is associated with structural aspects of one’s environment [18,19]. The sociodemographic make-up of residents in a neighborhood, which likely influences the social environment and resource distribution of neighborhoods, has also been associated with substance use behaviors [20,21]. There are numerous neighborhood measures that aim to capture social, structural, or sociodemographic characteristics of neighborhoods. These measures can be used to study how health behaviors differ between neighborhoods with high and low levels of specific characteristics.

This study aimed to understand how neighborhood-level factors were associated with the use of cigarettes, cannabis, and e-cigarettes in over 100,000 adults across Canada as part of the Canadian Partnership for Tomorrow’s Health (CanPath) cohort.

## Materials and Methods

### Study population

The data used in this study were collected as part of the CanPath cohort (101). CanPath is Canada’s largest longitudinal cohort, of over 330,000 Canadians across British Columbia, Alberta, Ontario, Quebec, and the Atlantic Provinces. Participants are being recruited in Saskatchewan and Manitoba but data was not available at the time of this study (2024). At CanPath study baseline (between 2009-2015), participants were recruited from across Canada. To be included in CanPath, participants had to be aged 30-74, reside in one of the participating Canadian provinces, and be able to complete questionnaires in English or French [22]. Participants and were asked to complete sociodemographic and health questionnaires and also consented to be contacted for future research and follow-up. A comprehensive follow-up questionnaire was then administered between 2016 and 2018. In total, 134,404 CanPath participants completed the first follow-up questionnaire, making it the most recent and comprehensive self-reported data on participants’ use of cigarettes, cannabis, and e-cigarettes. Access to the CanPath de-identified participant data was granted July 4, 2023. The study was conducted in accordance with the Declaration of Helsinki, and approved by the Joint Institutional Review Board BC Cancer/University of British Columbia (REB #H23-01508). Informed consent was obtained from all participants in the CanPath study.

Covariates used in regression models were drawn from the follow-up CanPath questionnaire when possible, to provide contemporaneous measures. Demographic and health related variables included age, sex, household income, marital status, perceived health status, and participants’ self-reported history of chronic diseases (cancer, diabetes, cardiovascular disease, and mental health conditions). Variables not queried in the follow-up questionnaire were obtained from the baseline questionnaire (2009-2015), including participant’s ethnicity, education, physical activity levels, body mass index (BMI), and fruit and vegetable consumption.

### Neighborhood measures

Data on neighborhood measures were generated through the Canadian Urban Environmental Health Research Consortium (CANUE) [23]. The six-digit postal code of CanPath participants who completed the follow-up questionnaire were linked to CANUE data to provide measures of the neighborhood where each participant resided: material deprivation, social deprivation, gentrification, and the Canadian Marginalization (CAN-Marg) indices. Neighborhood measures are at the level of dissemination area (DA). DAs cover all of Canada and are the smallest geographic area that standard census data is collected for, with the average population of DAs ranging from 400 to 700 [24]. The neighborhood measures available through CANUE were generated using data from the Statistics Canada census, primarily the census completed in 2016. Every five years Statistics Canada asks all Canadian households to respond to either a long form questionnaire (about 25% of households) or short form questionnaire. Questions in the census cover a range of primarily sociodemographic topics, such as education, income, religion, and housing. The census questionnaire can be completed online, through mail, or over the phone. The 2016 census had a response rate above 95% [25].

Material deprivation scores were generated for each DA in Canada based on the proportion of residents without a high school diploma, the employment to population ratio in the area, and the average personal income [26]. Using principal component analysis, a factor score was generated from the forementioned information and allowed the creation of population-weighted quintile values (1 to 5, least deprived to most deprived) for each DA. Social deprivation scores were generated for each DA in Canada based on the proportion of residents living alone, the proportion of residents that were separated, divorced, or widowed, and the proportion of single-parent families [26]. Quintile scores were generated in the same manner as for material deprivation. Material and social deprivation quintile scores scaled to the corresponding Canadian province of a neighborhood were used in the current study.

The Gentrification, Urban Interventions and Equity (GENUINE) tool was used to produce area-level measures of gentrification within Canada [27]. The tool uses multiple measures in the domains of income, housing, occupation, education, and age to produce measures of gentrification [27]. Information from the Statistics Canada Census in 2006 and 2016 was used to identify if gentrification occurred in census tracts between 2006 and 2016. A neighborhood was considered to be gentrifying if there was an increase in the pro-portion of residents with a university-level education, an increase in housing value, or an increase in rental costs, between 2006 and 2016, that was greater than that observed in the respective region to which a neighborhood belonged [27]. For the purpose of analysis, neighborhoods identified as non-gentrifiable (in 2006, the median neighborhood income was greater than the median income in the respective region) and stable (did not experience gentrification) were combined into one category that was compared against gentrifying neighborhoods.

CAN-Marg consists of four measures developed based on previous research on neighborhood deprivation and marginalization: household security (i.e. household and dwellings, residential instability), material resources (i.e. material deprivation), labor force participation (i.e. dependency), and immigration and visible minority (IVM) proportion (i.e. ethnic concentration) [28]. The measure of household security is developed using indicators of family structures, population demographics, home ownership, household density, and migration into the neighborhood [28]. The measure of material resources is similar to the aforementioned material deprivation score [28]. The measure of labor force participation captures the proportion of residents in a neighborhood who are over the age of 65 and the proportion of residents over the age of 15 who are unemployed [28]. The measure of IVM captures the proportion of residents in a neighborhood who are recent immigrants to Canada and/or who self-identify as visible minorities [28]. Factor scores, for each Canadian DA with census information, were computed from factor loadings and scores were used to divide DAs into quintiles for each measure. For this study, the CAN-Marg measures of household security, labor force participation, and IVM were studied since material resources is captured in the aforementioned material deprivation score.

### Study variables

Participants self-reported their cigarette use as either daily (at least one cigarette every day for the past 30 days), occasional (at least one cigarette in the past 30 days, but not every day), or none (did not smoke at all in the past 30 days). Those who reported never smoking cigarettes in their lifetime had their past 30-day cigarette use recorded as “none”. Participants who responded yes to ever using cannabis and yes to ever using cannabis once a month for over a year were asked about their cannabis use in the past 30 days. Participants reported the number the days they used cannabis in the past 30 days. Use was then categorized as none, infrequent use (1 to 5 days a month), occasional use (6 to 20 days a month), and frequent use (21 to 30 days a month). This categorization is similar to that of the American National Survey on Drug Use and Health [29,30]. About 50,500 participants did not respond to questions about their past 30-day cannabis use, as participants who responded that they had never used cannabis once a month for over a year and thus were not asked to report their current cannabis use patterns. Participants self-reported whether they had ever used an e-cigarette or “vaping” device. Those who responded yes were then asked if they had used an e-cigarette in the past 30 days. E-cigarette use was dichotomized as yes/no, as the number of days of use was not queried.

### Covariates

Covariates related to substance use and neighborhood measures based on evidence in existing literature were considered for inclusion in models. Categorization of covariates was based upon established thresholds or prior studies in CanPath [31,32]. Covariates included age, sex, region of residence, income, education, marital status, ethnicity), health perception, physical activity, fruit and vegetable consumption, BMI, and history of chronic conditions (cancer, diabetes, cardiovascular disease, and mental health conditions). Age was considered as a continuous variable. Sex was dichotomized as male/female. Study region reflected CanPath cohorts, with each participant belonging to either British Columbia, Alberta, Ontario, Quebec, or the Atlantic Provinces (Nova Scotia, New Brunswick, Prince Edward Island, and Newfoundland and Labrador). Annual household income was categorized <$25,000, $25,000 - $50,000, $50,000 -$75,000, $75,000 - $100,000, $100,000 - $150,000, and >$150,000. Education was categorized as high school or below, college level, or bachelor’s degree or higher. Marital status was dichotomized as single (including widows) or partnered. Ethnicity was dichotomized as white or non-white, as 93.2% of the sample was white. Health perception was categorized as excellent to very good, good, and fair to poor. Physical activity was categorized as low, moderate, or high, based on participants’ responses to the International, Physical Activity Questionnaire [33]. Fruit and vegetable consumption was the number of servings of fruit and vegetables consumed on a typical day. BMI was calculated from participants’ measured (if available) or self-reported height and weight and categorized as <25kg/m2, 25.0-29.9kg/m2, and >30kg/m2.

### Analytical sample

The analytical sample varied for the analysis of neighborhood measures and substance use depending on the variables under study. Participants were excluded from analysis if they had missing information on the substance use variable or the primary independent variable (neighborhood measure) for a particular model. Of the total 134,404 participants that completed the follow-up questionnaire, the number of participants eligible for final analysis varied from 128,240 to 51,639 depending on the missingness of the independent and dependent variable. Missing data for a particular substance use variable resulted in 6,390 participants excluded from analyses focused on cigarette use, 58,558 participants excluded from analyses focused on cannabis use, and 5,972 participants excluded from all analyses focused on e-cigarette use. The Multivariate Imputations by Chained Equation (MICE) package in R was used to perform multiple imputation for covariates with missing values. Age, sex, region, and health perception were correlated with missing values and were used as auxiliary variables in the multiple imputation to prevent/reduce bias in the generated datasets. Predictive mean matching within the MICE package was used to impute the missing values. As the percent of missing information for the imputed variables typically ranged between 30% and 40%, 30 imputed datasets were generated, with 20 iterations required to achieve convergence.

### Statistical analysis

Means and standard deviations were reported for numeric variables and percentages were reported for categorical variables. Separate regression models were built to study how cigarette, cannabis, and e-cigarette use were associated with each neighborhood measure. Multinomial models were built to study how cannabis use and cigarette use were associated with the neighborhood measures being explored in this study as estimated via odds ratios (ORs). For e-cigarette use, logistic regression models were built to study as-sociations with neighborhood measures via ORs. Model assumptions were examined through residual plots.

Model 1 was unadjusted. Model 2 adjusted for sex, age, and regional cohort. For-ward stepwise regression was then performed for all other covariates. Variables that changed the estimate by ∼10% and were statistically significant (p-value <0.05) remained in the adjusted model. Interactions between the neighborhood measures and sex were examined for each model. For all the models examining neighborhood-level variables, the interaction between the neighborhood measure and region was also examined. Interactions with other covariates were explored in some models based on subject area knowledge. For example, interactions between gentrification and household income were tested in models examining associations between substance use and gentrification, although no interactions at the neighborhood level were subsequently found to be significant.

All analyses and data cleaning were conducted in R Studio (R version 4.2.3) [34]. The nnet package was used to analyze multinomial models [34]. A confidence level of 95% was used as a threshold for statistical significance.

## Results

### Population descriptives

The demographic, lifestyle, and health characteristics of all CanPath participants who completed the follow up survey are presented in Table 1. The mean age of participants was 60y, and 65.1% of participants were female. The majority of participants had a household income >$50,000 and 83.1% of participants were university or college educated. When asked to rate their health, 82.9% of participants rated their health as good to excellent. Demographic, lifestyle, and health characteristics are also reported within categories of cigarette, cannabis, and e-cigarette use in the supplemental Tables S1 to S3.

**Table 1.**
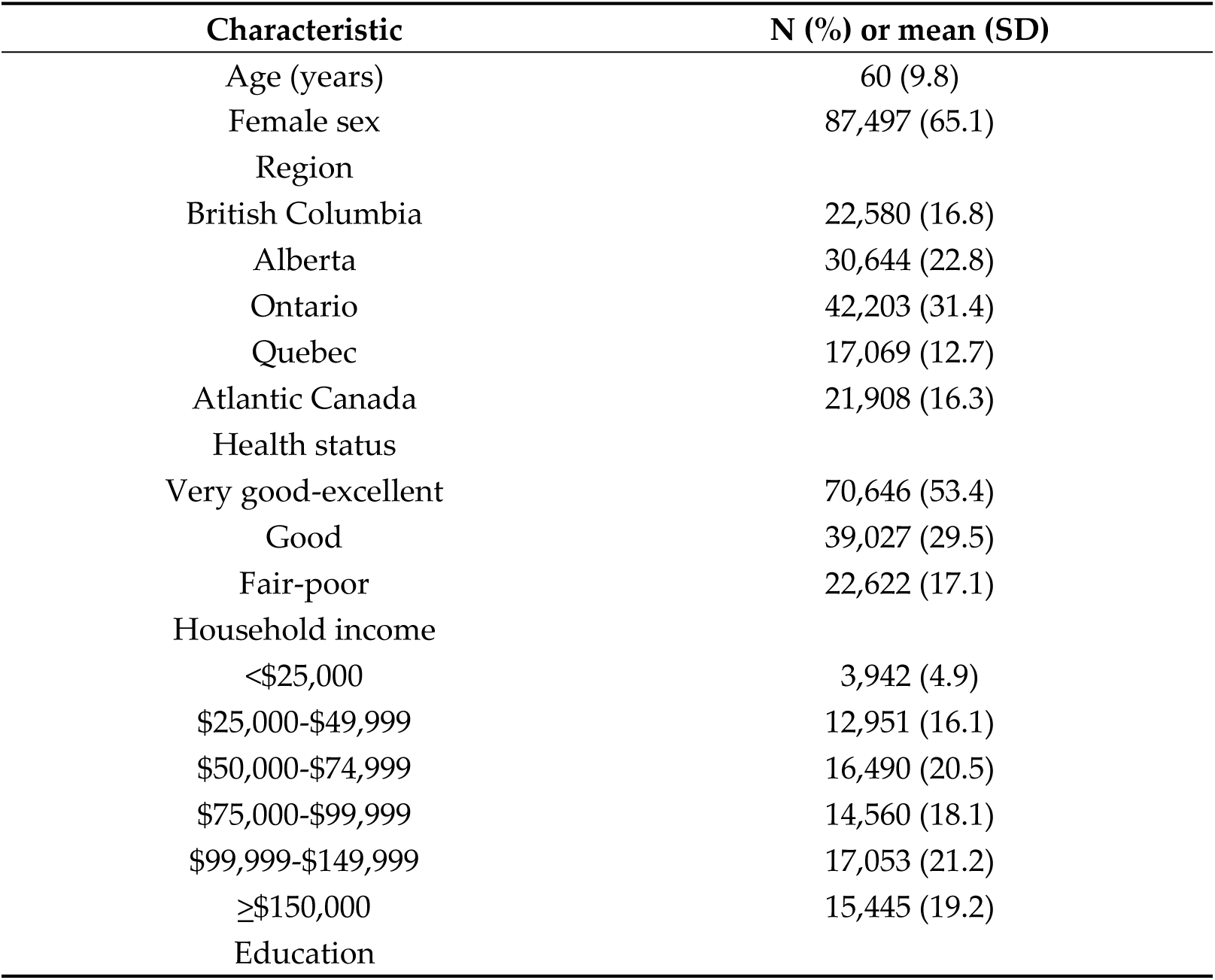
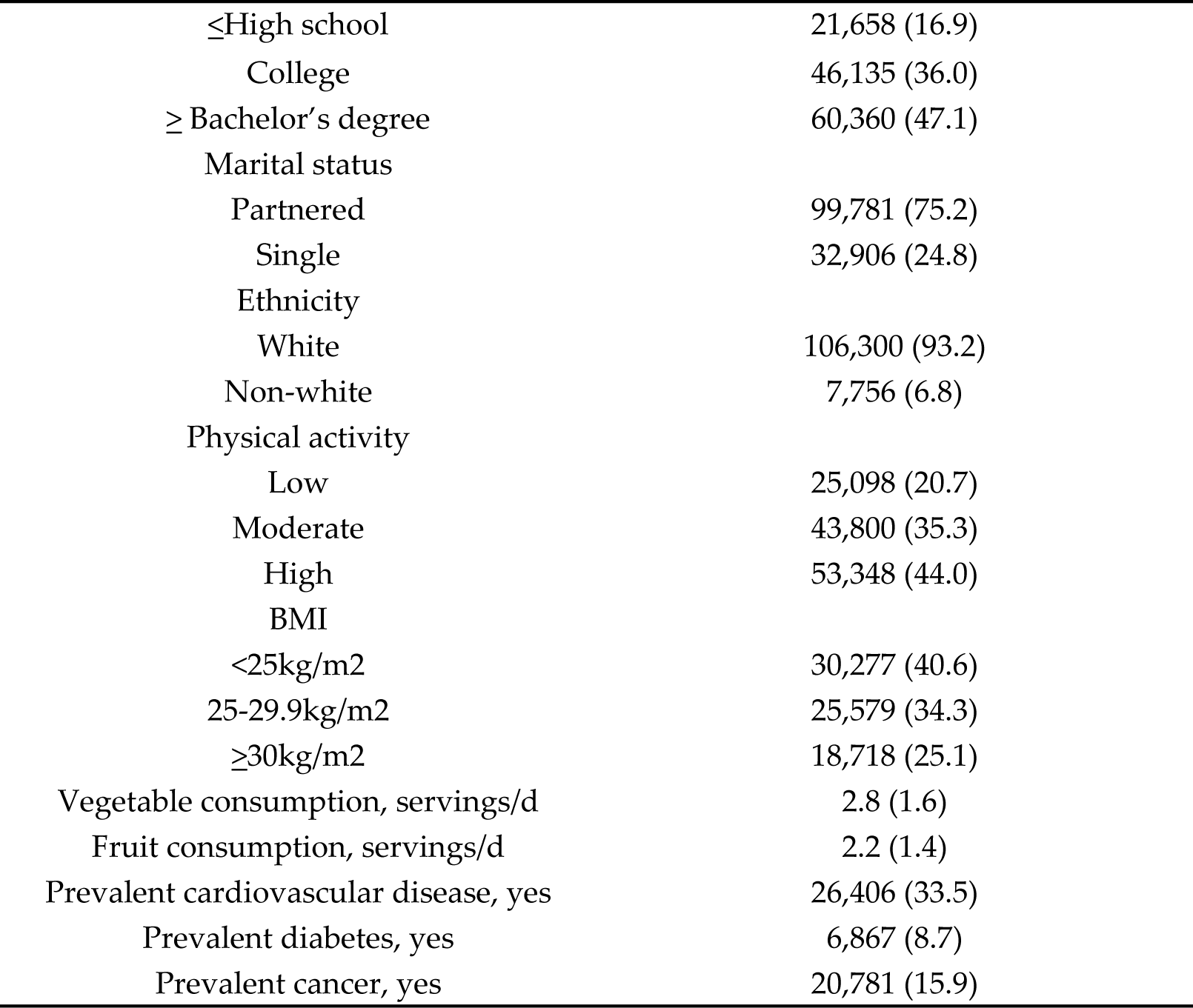
Characteristics of CanPath participants (N=134,404) who completed the follow-up. The prevalence of cigarette, cannabis, and e-cigarette use for CanPath participants who completed the follow-up questionnaire is presented in Table 2. Only 4.2% of participants smoked cigarettes daily, and only 2.3% used cannabis frequently. In the past 30-days, only 1% of participants reported cannabis use.

**Table 2.**
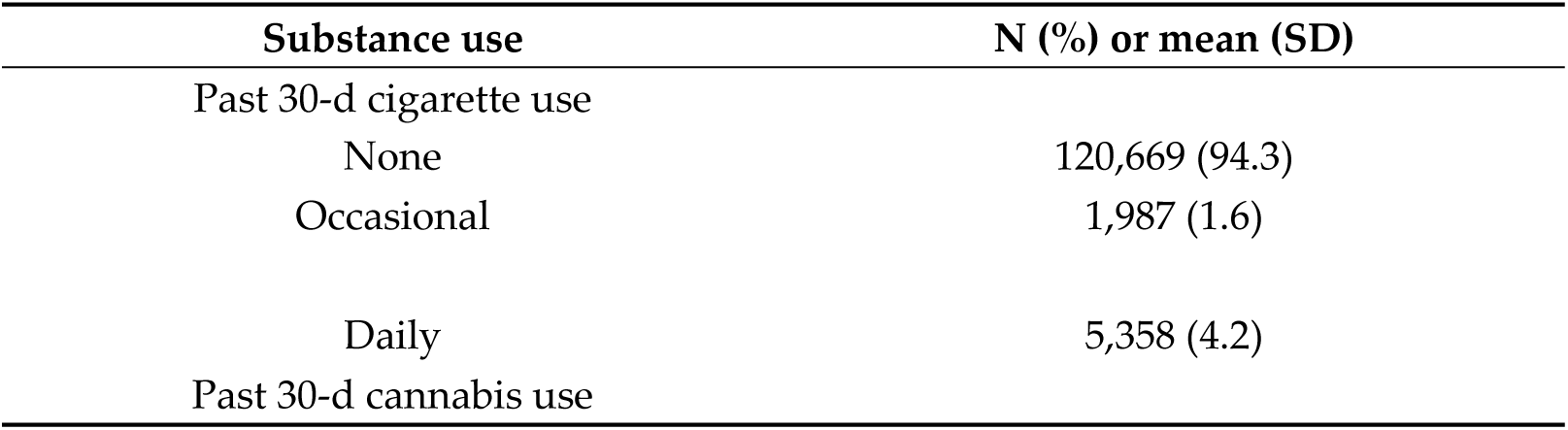
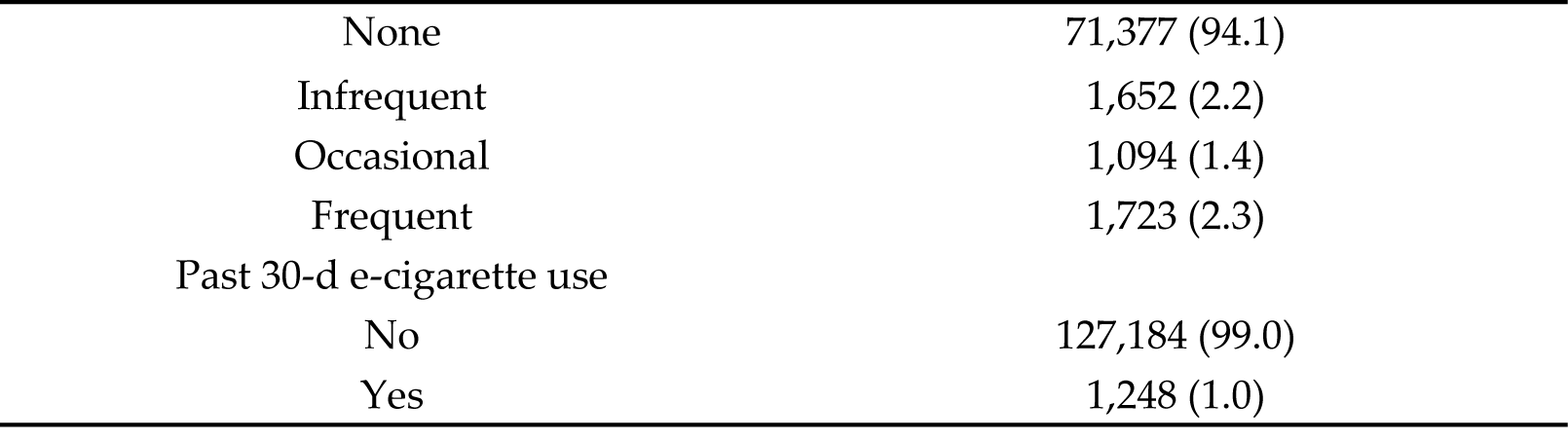
Descriptives of substance use for all. CanPath participants who completed the follow-up questionnaire.

Descriptives of neighborhood measures by categories of substance use for all participants who completed the follow-up questionnaire are presented in Table S4-S6. More participants lived in neighbourhoods in the lowest two quintiles of material deprivation (least deprived) than in the highest two quintiles of material deprivation (most deprived), 57.5% vs. 23.6%, respectively. Similarly, for the CAN-Marg index of material resources, with 59.8% and 21.9% of participants living in neighbourhoods in bottom two and top two quintiles, respectively. Between 17.9% and 1.6% of participants lived in neighbourhoods that had experienced gentrification between 2006 and 2016 depending on the measure of gentrification used.

### Cigarette smoking

Associations between cigarette smoking and neighborhood measures are shown in Table 3. In model 2, the odds of smoking cigarettes occasionally were 32% (OR = 1.32, 95% CI = 1.11, 1.56) higher for participants living in the most deprived neighborhoods (fifth quintile) compared to those living in the least deprived neighborhoods (first quintile). After adjusting for confounders, the odds ratio of smoking cigarettes daily was higher in the second to fifth quintile of material deprivation compared to the least deprived quintile. For example, OR= 2.00, 95% CI = 1.80, 2.22 (model 2), for participants living in the most deprived neighborhoods versus those living in the least deprived neighbourhoods. For social deprivation, the odds of occasionally smoking cigarettes were 25% (OR = 1.25, CI =1.07, 1.46, model 2) and 36% (OR = 1.36, CI = 1.16, 1.59, model 2) higher for participants living in neighborhoods in the fourth and fifth quintile of social deprivation, respectively, compared to participants living in neighborhoods in the first quintile. The odds of daily cigarette smoking were higher for participants living in neighborhoods from the second to fifth quintile of social deprivation than for participants living in neighborhoods in the first quintile in the unadjusted and adjusted models.

**Table 3.**
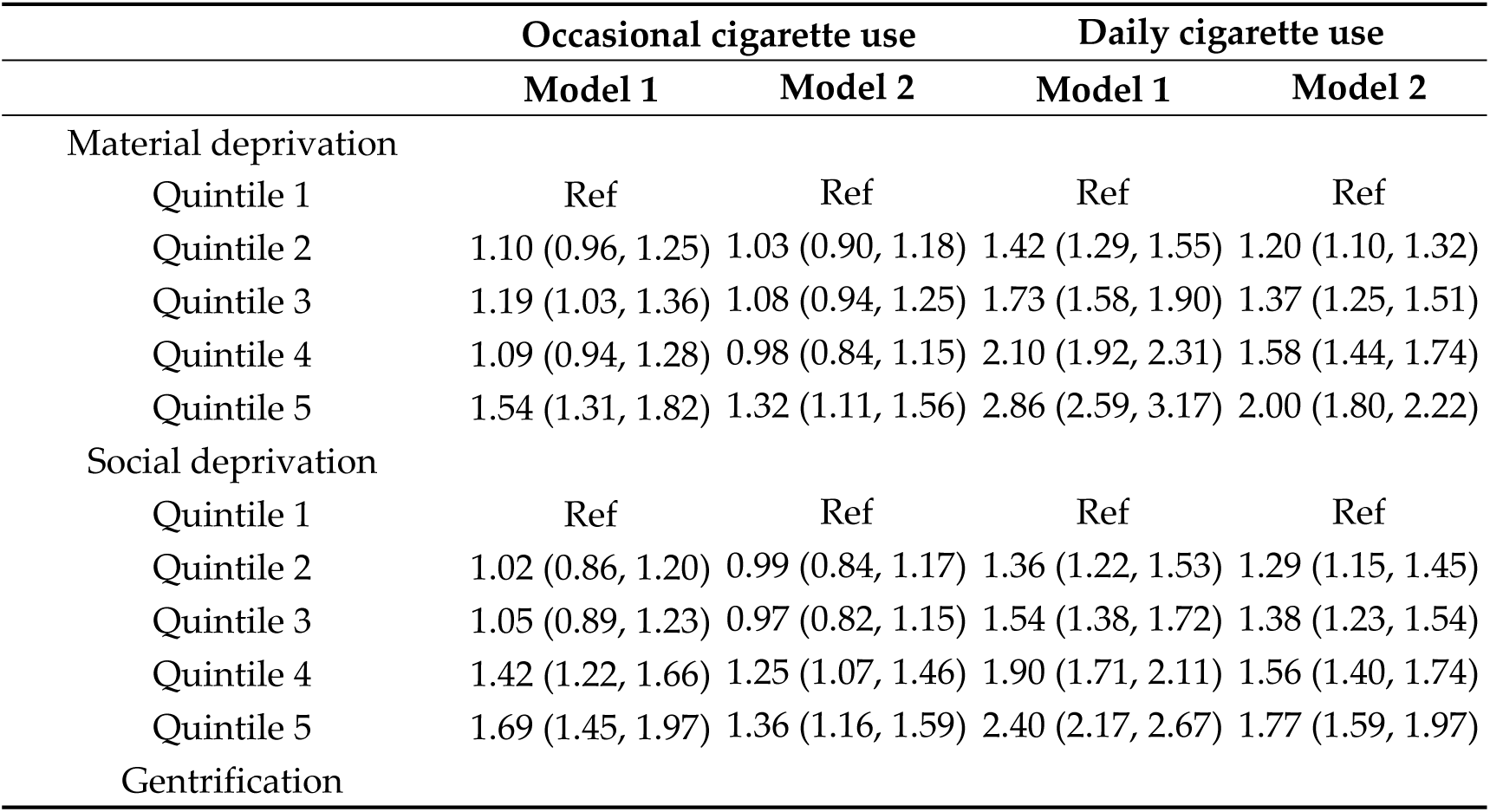
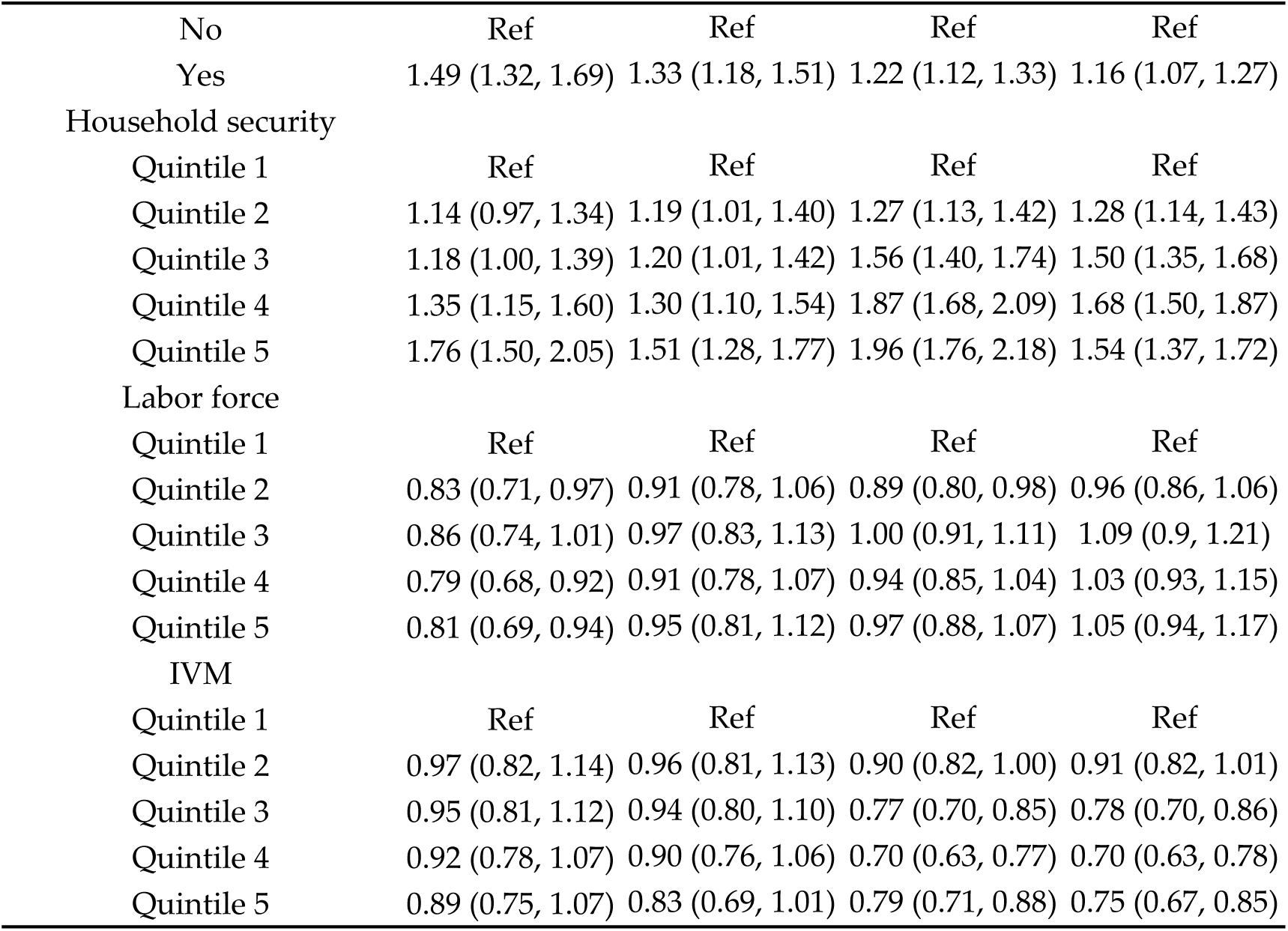
Odds ratios of cigarette use (95% Cis) by neighborhood characteristics. Quintile 1 represents least deprived (material and social deprivation), most secure, most employment, and least IVM residents. Model 1was unadjusted. Model 2 adjusted for age, sex, and region and covariates that differed depending on the neighborhood measure due to stepwise covariate selection. Material deprivation adjusted for health status, and education, social deprivation, gentrification, labor force and IVM adjusted for income and marital status. Reference is no cigarette use. IVM: immigration and visible minority.

In model 1 and 2, the odds of smoking cigarettes occasionally or daily were higher for participants living in gentrified neighborhoods than for participants living in non-gentrified neighborhoods. For instance, OR=1.33 (95% CI 1.18, 1.51) for occasional cigarette use in gentrified neighborhoods compared to non-gentrified. The odds of occasional cigarette smoking was lower among participants living in neighborhoods with lower labour force participation (quintiles 4 and 5), but estimates were attenuated in model 2. No associations were observed between daily cigarette use and labor force. Both model 1 and 2 found that the odds of smoking cigarettes daily were lower for participants who lived in neighborhoods in quintiles 3-5of IVM relative to quintile 1. For example, adjusted OR = 0.75 (95% CI = 0.67, 0.85) for participants living in neighborhoods in quintile five versus quintile one of IVM. There were no associations between occasional smoking and neighborhood IVM.

### Cannabis use

Associations between cannabis use and neighborhood factors are shown in Table 4. There were no significant associations between infrequently or occasionally using cannabis and living in a materially deprived neighborhood. After adjusting for confounders (model 2), the odds of frequently using cannabis were 24% (OR = 1.24, 95% CI = 1.06, 1.46) and 48% (OR = 1.48, 95% CI = 1.24, 1.77) higher for participants living in neighborhoods in the fourth and fifth quintiles of material deprivation, compared to participants living in neighborhoods in the first quintile. After adjusting for confounders, the odds of infrequently, occasionally, and frequently using cannabis were 94% (OR = 1.94, 95% CI = 1.63, 2.31), 117% (OR = 2.17, 95% CI = 1.74, 2.71), and 196% (OR = 2.96, 95% CI = 2.43, 3.60) higher, respectively, for participants living in neighborhoods in the fifth quintile of social deprivation than for participants living in neighborhoods in the first quintile. The odds of infrequent, occasional and frequent cannabis use were all higher for participants living in gentrified neighborhoods than for participants living in non-gentrified neighbourhoods (e.g. frequent use: OR = 2.04, 95% CI = 1.79, 2.33, model 2). Th

**Table 4.** Odds ratios of cannabis use (95% CIs) by neighborhood characteristics. Quintile 1 represents least deprived (material and social deprivation), most secure, most employment, and least IVM residents. 2Model 1 was unadjusted. Model 2 adjusted for age, sex, and region and covariates that differed depending on the neighborhood measure due to stepwise covariate selection. Material deprivation adjusted for health, income, education and ethnicity, social deprivation: marital status, gentrification: education and marital status, household security, labor force and IVM: income and marital status. Reference is no cannabis use. IVM: immigration and visible minority.

|  | Infrequent |  | Occasional |  | Frequent |  |
| --- | --- | --- | --- | --- | --- | --- |
|  | Model 1 | Model 2 | Model 1 | Model 2 | Model 1 | Model 2 |
| Material deprivation |  |  |  |  |  |  |
| Quintile 1 | Ref | Ref | Ref | Ref | Ref | Ref |
| Quintile 2 | 0.96 (0.84, 1.11) | 0.89 (0.78, 1.03) | 0.91 (0.76, 1.09) | 0.83 (0.69, 0.99) | 1.03 (0.88, 1.20) | 0.87 (0.74, 1.02) |
| Quintile 3 | 0.86 (0.73, 1.00) | 0.79 (0.68, 0.93) | 0.99 (0.81, 1.19) | 0.88 (0.73, 1.07) | 1.28 (1.09, 1.49) | 1.03 (0.87, 1.20) |
| Quintile 4 | 0.96 (0.81, 1.13) | 0.89 (0.75, 1.05) | 1.20 (0.99, 1.45) | 1.07 (0.87, 1.30) | 1.61 (1.37, 1.88) | 1.24 (1.06, 1.46) |
| Quintile 5 | 1.01 (0.83, 1.23) | 0.91 (0.75, 1.11) | 1.10 (0.87, 1.40) | 0.94 (0.74, 1.20) | 2.10 (1.78, 2.49) | 1.48 (1.24, 1.77) |
| Social deprivation |  |  |  |  |  |  |
| Quintile 1 | Ref | Ref | Ref | Ref | Ref | Ref |
| Quintile 2 | 1.05 (0.87, 1.27) | 1.08 (0.89, 1.31) | 1.13 (0.88, 1.44) | 1.16 (0.91, 1.48) | 1.74 (1.41, 2.15) | 1.75 (1.42, 2.16) |
| Quintile 3 | 1.48 (1.24, 1.76) | 1.51 (1.26, 1.80) | 1.56 (1.25, 1.95) | 1.60 (1.28, 2.00) | 1.94 (1.58, 2.38) | 1.90 (1.55, 2.34) |
| Quintile 4 | 1.50 (1.26, 1.79) | 1.50 (1.25, 1.79) | 1.76 (1.41, 2.19) | 1.76 (1.41, 2.20) | 2.74 (2.26, 3.33) | 2.55 (2.09, 3.10) |
| Quintile 5 | 2.06 (1.73, 2.44) | 1.94 (1.63, 2.31) | 2.29 (1.84, 2.84) | 2.17 (1.74, 2.71) | 3.55 (2.93, 4.30) | 2.96 (2.43, 3.60) |
| Gentrification |  |  |  |  |  |  |
| No | Ref | Ref | Ref | Ref | Ref | Ref |
| Yes | 2.08 (1.82, 2.36) | 1.92 (1.68, 2.19) | 1.99 (1.70, 2.34) | 1.85 (1.57, 2.17) | 2.27 (2.00, 2.58) | 2.04 (1.79, 2.33) |
| Household security |  |  |  |  |  |  |
| Quintile 1 | Ref | Ref | Ref | Ref | Ref | Ref |
| Quintile 2 | 1.16 (0.96, 1.41) | 1.20 (0.99, 1.45) | 1.67 (1.31, 2.14) | 1.74 (1.36, 2.22) | 1.46 (1.19, 1.78) | 1.48 (1.21, 1.82) |
| Quintile 3 | 1.50 (1.25, 1.80) | 1.53 (1.27, 1.84) | 1.94 (1.53, 2.47) | 2.00 (1.57, 2.54) | 2.12 (1.75, 2.57) | 2.09 (1.72, 2.53) |
| Quintile 4 | 1.72 (1.43, 2.07) | 1.71 (1.42, 2.06) | 2.37 (1.87, 3.01) | 2.39 (1.87, 3.04) | 2.29 (1.89, 2.78) | 2.13 (1.75, 2.59) |
| Quintile 5 | 2.30 (1.93, 2.75) | 2.14 (1.79, 2.57) | 2.96 (2.35, 3.74) | 2.80 (2.20, 3.55) | 3.22 (2.67, 3.87) | 2.62 (2.16, 3.17) |
| Labor force |  |  |  |  |  |  |
| Quintile 1 | Ref | Ref | Ref | Ref | Ref | Ref |
| Quintile 2 | 0.89 (0.75, 1.05) | 0.96 (0.81, 1.14) | 0.83 (0.68, 1.02) | 0.90 (0.73, 1.11) | 1.01 (0.86, 1.19) | 1.11 (0.94, 1.32) |
| Quintile 3 | 0.84 (0.71, 0.99) | 0.95 (0.80, 1.13) | 0.89 (0.73, 1.09) | 1.02 (0.83, 1.25) | 0.88 (0.74, 1.04) | 1.02 (0.86, 1.22) |
| Quintile 4 | 0.83 (0.70, 0.98) | 0.99 (0.83, 1.18) | 0.82 (0.67, 1.01) | 0.98 (0.80, 1.21) | 0.80 (0.68, 0.95) | 0.99 (0.83, 1.18) |
| Quintile 5 | 0.77 (0.65, 0.92) | 0.99 (0.82, 1.18) | 0.73 (0.59, 0.91) | 0.94 (0.75, 1.17) | 0.90 (0.76, 1.07) | 1.16 (0.98, 1.39) |
| IVM |  |  |  |  |  |  |
| Quintile 1 | Ref | Ref | Ref | Ref | Ref | Ref |
| Quintile 2 | 1.15 (0.96, 1.37) | 1.09 (0.91, 1.30) | 0.83 (0.67, 1.02) | 0.78 (0.63, 0.97) | 0.94 (0.79, 1.11) | 0.91 (0.76, 1.08) |
| Quintile 3 | 1.06 (0.89, 1.26) | 0.95 (0.79, 1.13) | 0.93 (0.76, 1.13) | 0.82 (0.67, 1.01) | 0.80 (0.68, 0.95) | 0.73 (0.61, 0.87) |
| Quintile 4 | 0.97 (0.81, 1.16) | 0.80 (0.67, 0.97) | 0.84 (0.68, 1.03) | 0.68 (0.55, 0.85) | 0.83 (0.71, 0.98) | 0.69 (0.58, 0.82) |
| Quintile 5 | 0.81 (0.66, 0.99) | 0.60 (0.49, 0.74) | 0.64 (0.50, 0.81) | 0.46 (0.36, 0.60) | 0.75 (0.62, 0.91) | 0.54 (0.44, 0.66) |

The odds of occasionally and frequently using cannabis were higher for participants living in neighborhoods in the second to fifth quintile of household security compared to participants living in neighborhoods in the first quintile, in both model 1 and 2. Associations between neighborhood labor force and cannabis use was not significant with adjustment for covariates in model 2. The odds of frequently using cannabis were 46% lower (OR = 0.54, 95% CI = 0.44, 0.66) for participants living in neighborhoods in the fifth quintile of IVM than for participants living in neighborhoods in the first quintile. The odds of frequent cannabis use were also lower for participants living in neighbourhoods in fourth quintile and third quintile of IVM. The odds of infrequent cannabis use were lower in the adjusted model for participants living in neighborhoods in the fifth and fourth quintile of IVM.

### E-cigarette use

After adjusting for covariates, the odds of past 30-day e-cigarette use were higher for participants who lived in neighborhoods in the fourth and fifth quintile of material deprivation (quintile 5, model 2: OR = 1.42, 95% CI = 1.15, 1.76). The odds of e-cigarettes use were higher in quintile 4 and quintile 5 of socially deprived neighborhoods in model 1 and 2: quintile 4: OR = 1.28, 95% CI = 1.04, 1.58), and quintile 5: OR = 1.52, 95% CI =1.24, 1.87. The odds of e-cigarette use was also higher in gentrified neighborhoods, but only in model 1. With respect to household security, the odds of e-cigarette use were 36% (OR = 1.36, 95% CI = 1.10, 1.68) and 30% (OR = 1.30, 95% CI = 1.05, 1.61) higher for participants living in neighborhoods in the fourth and fifth quintile of household security (model 2). Associations between e-cigarette use and labor force were non-significant in model 2. For IVM, the odds of e-cigarette use were 28% lower (OR =0.72, 95% CI = 0.56, 0.93) for participants living in neighborhoods in the fifth quintile of IVM than for participants living in neighborhoods in the first quintile.

**Table 5.**
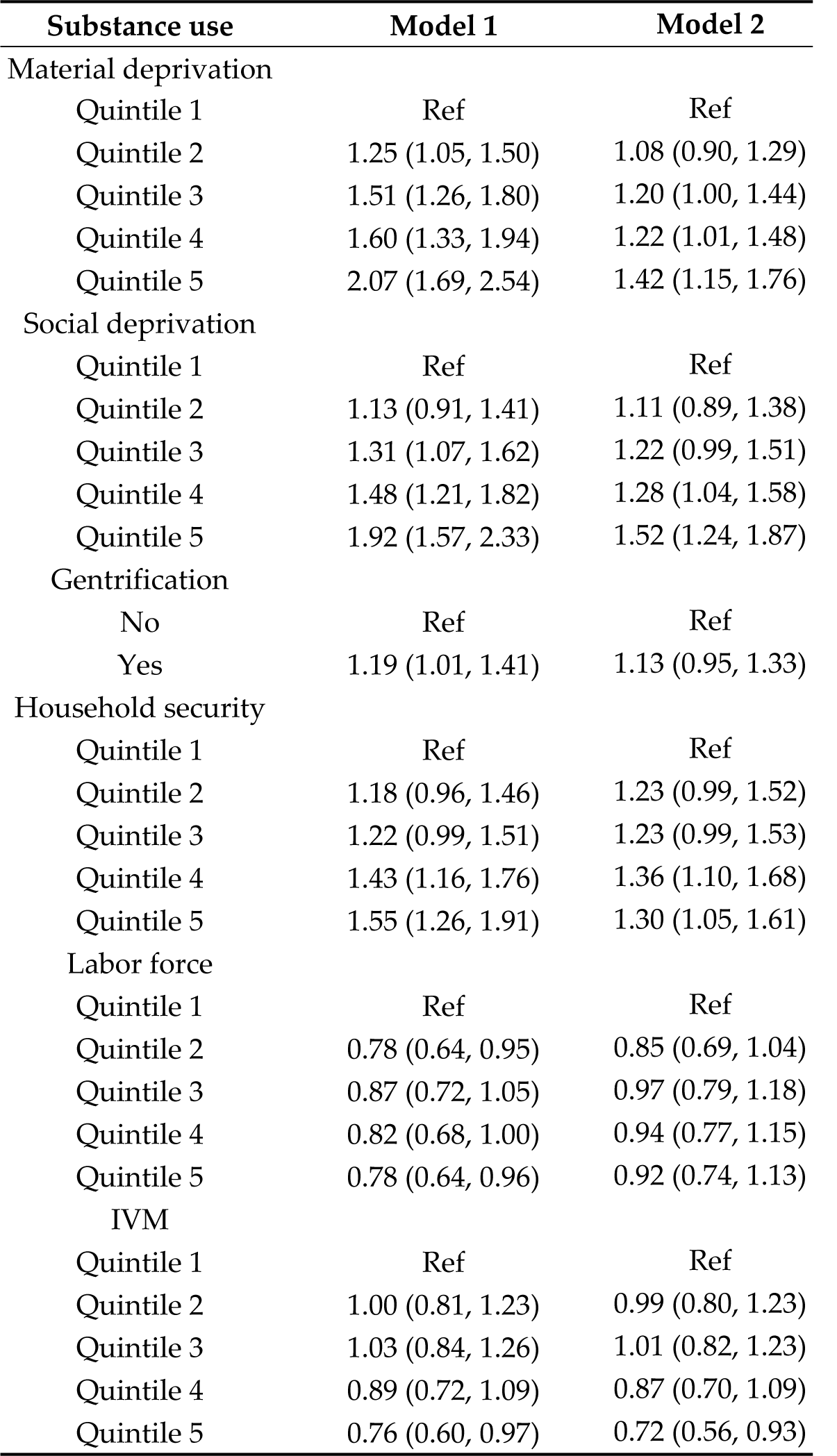
Odds ratios of e-cigarette use (95% CIs) by neighborhood characteristics. Quintile 1 represents least deprived (material and social deprivation), most secure, most employment, and least IVM residents. 2Model 1 was unadjusted. Model 2 adjusted for age, sex, and region and covariates that differed depending on the neighborhood measure due to stepwise covariate selection. Material deprivation adjusted for health, income and education, social deprivation, household security, labor force and IVM: income and marital status, gentrification: income, education and marital status. Reference is no e-cigarette use. IVM: immigration and visible minority.

## Discussion

The current study provides estimates of the prevalence of use of legal substances use among adults (age 30 to 85) from diverse geographical areas in Canada. Aside from surveys performed by Statistics Canada, there are limited contemporary estimates of the prevalence of cannabis and e-cigarette use in Canada. Data on the prevalence of the use of these substances in a large pan-Canadian adult population is important for surveillance and monitoring. This is also one of the few studies to examine multiple neighbor-hood-level risk factors for substance use in Canada. Notable findings include robust positive associations between neighborhood material and social deprivation with occasional and daily cigarette smoking, e-cigarette use and frequent cannabis use.

Previous research on neighborhood material and social deprivation and substance use is relatively scarce and few studies use indicators of deprivation that are comparable to the ones used in the current study. A study in the U.S. found that greater neighborhood socioeconomic disadvantage was associated with a greater likelihood of cigarette smoking [35]. Similarly, another study in the U.S. found that participants living in neighborhoods with high levels of socioeconomic disadvantage were more likely to smoke cigarettes [20]. The measures of socioeconomic disadvantage had similarities to the measure of material deprivation used in the current study, with overlapping metrics being the education and income level of neighborhood residents. Research in the U.S. has also found an association between living in neighborhoods with greater social capital, assessed based on individual perceptions of the neighborhood social environment, and less cigarette and alcohol use [20]. This agrees with our findings of greater use of these substance for participants living in more socially deprived neighborhoods. Another study in the U.S. found that neighborhood disadvantage was associated with illicit drug use, which included cannabis use [21]. In a secondary analysis, the previously mentioned study found that the relationship between neighborhood disadvantage and illicit drug use largely acted through individuals’ exposure to social stressors and feelings of psychological distress [21].

One plausible explanation for these observed associations between substance use and neighborhood factors draws upon the structural hypothesis [14]. This hypothesis would suggest that health behaviors, in this case substance use behaviors, differ between more and less deprived neighborhoods due to differences in social norms and the built environment [14]. There may be differences in the social environment of neighborhoods with greater material and social deprivation, which could shape the attitudes and beliefs that residents adopt and affect the supports, resources, and information available to residents [36]. Built structures within neighborhoods with greater material and social deprivation could also differ, which may influence how public resources (e.g. community centers, parks, etc.), retail spaces (including those for cigarettes, cannabis, and e-cigarettes), and other institutions and organizations are accessible to neighborhood residents [14]. For example, more cannabis retailers are located in socially deprived neighborhoods (quintiles 4 and 5) than in non-socially deprived neighborhoods in Canada [37]. Although the private sector may also themselves influence the social, physical and cultural environments, also known as commercial determinants of health [38].

This studies’ finding that living in a gentrified neighborhood is associated with higher odds of cigarette, cannabis, and e-cigarette use is novel within the Canadian context. Qualitative research has found that living in a gentrified neighborhood can impact health by causing housing and financial insecurity, socio-cultural displacement, and a loss of amenities for residents [39]. Those living in gentrified neighborhoods are also more likely to experience psychological stress [39]. These negative effects of gentrification are typically observed in marginalized groups; residents with low-income, racial and ethnic minorities, and older residents [39].

Few previous studies have examined associations between neighborhood household security and health behaviors. One study in the U.S. found that neighborhood residential stability, a measure similar to household security, was not associated with cigarette or alcohol use [20]. A study in Ontario found that neighborhood household security was positively associated with avoidable mortality, which included premature deaths from health conditions linked to substance use [40]. The measure of household security in this study reflects neighborhood cohesion and support using multiple indicators of household security. It is therefore difficult to make assumptions about what factors may be driving the associations with substance use that were observed in the current study.

Living in a neighborhood with a higher proportion of residents who are recent immigrants and/or visible minorities was associated with lower odds of cigarette, can-nabis, and e-cigarette use. Previous research has found that immigrants to Canada have lower levels or alcohol, cigarette, and cannabis use compared to non-immigrants [41]. The findings of this study support the “healthy immigrant effect”, which suggests recent immigrants to Canada have better health than those born in Canada [42]. Using so-cial-interactive theory, it is possible that living in a neighborhood with a higher pro-portion of immigrants may lead individuals to have greater social contact with people who are not often using substances, resulting in lower substance use behaviors among all neighborhood residents [36]. There is no research on immigrant or visible minority populations and e-cigarette use in Canada. Evidence from the current study suggests that trends of e-cigarette use among recent immigrants to Canada may be similar to those observed for cigarette and cannabis use.

In contrast to the other neighborhood factors, the proportion of residents in a neighborhood participating in the labor force was not associated with cigarette, cannabis, or e-cigarette use. As our study looked at general non-participation in the labor force, it is possible there was no observed association with substance use because of the variety of reasons people may be unemployed. Future research that incorporates different reasons for unemployment into analyses could be important to elucidate the relationship between neighborhood labor force and substance use.

A strength of this study is the large number of individuals from diverse geo-graphical areas in Canada that differ with respect to demographics, population density and structural factors. There is very little research using similar data in Canada and studies from the U.S. tend to focus on select cities or regions. Another strength is the information on a wide range of demographic, lifestyle, and health characteristics for participants. This allowed for consideration of multiple potential confounders in models. The main strength of this study was our ability to connect data on participants’ substance use behaviors to multiple measures of their neighborhood environment. Compared to individual-level risk factors, our understanding of how neighborhood characteristics are associated with substance use behaviors is limited. Multiple studies in Canada have however observed greater mortality risk for individuals living in more materially and socially deprived neighborhoods [43,44], although reasons why were unclear. Findings from the current study suggest that increased substance use could be a potential explanation.

A limitation of this study is the cross-sectional design. Although, it is unlikely that an individual’s substance use patterns would lead them to live in a certain neighborhood [14]. Another limitation is that the CanPath cohort is not representative of the Canadian population. Selection bias could impact the generalizability of our findings. Compared to the Canadian population, our study population contained a higher proportion of females (65.1% vs. 50.3%), was older (mean age of 60 vs 41), and was more likely to be white (93.2% vs. 73.5%) than the general Canadian population [45]. Compared to the 2020 Statistics Canada census, study participants also had higher incomes and were more likely to have a bachelor’s degree or higher [45]. There was no representation of Canadians from Saskatchewan, Manitoba, Nunavut, Yukon, or the Northwest Territories in the CanPath study at the time of the follow-up questionnaire. As a result of the CanPath study population, the use of cigarettes, cannabis, and e-cigarettes was lower than in the Canadian general population, which may have also reduced the power and biased findings towards the null. Another limitation is that neighborhood measures were generated using 2016 census data and substance use behaviors were reported in the 2016-2018 follow-up questionnaire. The potential disconnect could have resulted in misclassification of participants’ neighborhood environment exposures at the time when they reported on their current substance use patterns, although neighborhoods are considered to be relatively stable over time [26,46]. The numerous analyses examining associations between neighborhood factors and substance use increased the likelihood of spurious associations being found. Statistical approaches to adjust for multiple comparisons through approaches like the Bonferroni correction and the q-value were not performed due to the exploratory nature of the study, particularly with respect to neighborhood-level factors and substance use.

## Conclusions

As we provide evidence that neighborhood characteristics are associated with substance use, it follows that interventions aimed at the neighborhood or community level may also be effective for reducing or preventing substance use. Specifically, interventions aimed at reducing or preventing the use of these substances could be directed at individuals living in more materially or socially deprived neighborhoods. The evidence presented in this study could be used to inform policy makers and urban planners in Canada about neighborhood gentrification and potentially inform zoning, or neighborhood development. These findings also provide rational for researchers to conduct further studies on gentrification and health in Canada, ideally research that advances our understanding of the scope of the issue and which groups or regions are most affected. Collectively, the findings could help identifying populations who are at an increased risk of using substances and ensure accessibility of resources.

## Data Availability

Software code is available from researchers upon reasonable re-quest. Code used to derive variables is returned to the CanPath as per data access requirements. The datasets analyzed in this study were made available by the Canadian Partnership for To-morrow’s Health (CanPath) and are available on reasonable request and with approval from the CanPath.

## Supplementary Materials

Table S1: Demographic, lifestyle, and health characteristics for participants who did not smoke (none), smoked cigarettes occasionally or daily; Table S2: Demographic, lifestyle, and health characteristics for participants who used cannabis frequently, occasionally, infrequently, or not at all during the past 30 days Table S3: Demographic, lifestyle, and health characteristics for participants who reported using or not using an e-cigarette in the past 30 days. Table S4: Descriptive statistics, N (%), for the prevalence of neighborhood factors within levels of cigarette use. Table S5: Descriptive statistics, N (%), for the prevalence of neighborhood factors within levels of past 30-day cannabis use. Table S6: Descriptive statistics, N (%), for the prevalence of neighborhood factors among participants who did and did not use an e-cigarette in the past 30-days.

## Author Contributions

Conceptualization, TFW; methodology, TFW, RAM.; formal analysis, TFW.; investigation, TFW; resources, RAM, TJBD.; writing—original draft preparation, TFW, RAM.; writing—review and editing, CP, TJBD.; supervision, RAM, TJBD, CP. All authors have read and agreed to the published version of the manuscript.

## Data Availability Statement

eth

## Acknowledgments

A version of this work was published as part of TFW’s Master’s thesis. Doi: 10.14288/1.0442427. The Canadian Partnership for Tomorrow’s Health (CanPath) research is only possible with the commitment of its research participants, its staff, and its funders. The data used in this research were made available by CanPath and BC Generations Project, Alberta’s To-morrow Project, the Ontario Health Study, CARTaGENE, and the Atlantic Partnership for Tomorrow’s Health.

